# VALIDATION OF A SALIVA-BASED TEST FOR THE MOLECULAR DIAGNOSIS OF SARS-CoV-2 INFECTION

**DOI:** 10.1101/2021.09.10.21263072

**Authors:** Michela Bulfoni, Emanuela Sozio, Barbara Marcon, Maria De Martino, Daniela Cesselli, Chiara De Carlo, Romina Martinella, Angelica Migotti, Eleonora Vania, Agnese Zanus-Fortes, Jessica De Piero, Emanuele Nencioni, Carlo Tascini, Miriam Isola, Francesco Curcio

## Abstract

**Background:** Since the beginning of the pandemic, clinicians and researchers have been searching for alternative tests to improve screening and diagnosis of SARS-CoV-2 infection (*Y. Yang et al*., *medRxiv 2020; W. Wang et al*., *2020*.*3786; A Senok et al*., *Infect Drug Resist 2020)*. Currently, the gold standard for virus identification is the nasopharyngeal (NP) swab (N. Sethuraman et al., JAMA 2020; A.J. Jamal et al Clinical Infect Disease 2021). Saliva samples, however, offer clear practical and logistical advantages (*K*.*K*.*W To et al, Clinical Microb and Infect; A*.*L. Wylle et al. N Engl J Med 2020; N. Matic et al, Eur J Clin 2021*) but due to lack of collection, transport, and storage solutions, high-throughput saliva-based laboratory tests are difficult to scale up as a screening or diagnostic tool (D. *Esser et al*., *Biomark Insights 2008; E. Kaufman et al*., *Crit Rev Oral Biol Med2002)*. With this study, we aimed to validate an intra-laboratory molecular detection method for SARS-CoV-2 on saliva samples collected in a new storage saline solution, comparing the results to NP swabs to determine the difference in sensitivity between the two tests.

**Methods:** In this study, 156 patients (cases) and 1005 asymptomatic subjects (controls) were enrolled and tested simultaneously for the detection of the SARS-CoV-2 viral genome by RT-PCR on both NP swab and saliva samples. Saliva samples were collected in a preservative and inhibiting saline solution (Biofarma Srl). Internal method validation was performed to standardize the entire workflow for saliva samples.

**Results:** The identification of SARS-CoV-2 conducted on saliva samples showed a clinical sensitivity of 95.1% and specificity of 97.8% compared to NP swabs. The positive predictive value (PPV) was 81% while the negative predictive value (NPV) was 99.5%. Test concordance was 97.6% (Cohen’s Kappa=0.86; 95% CI 0.81-0.91). The LoD of the test was 5 viral copies for both samples.

**Conclusions:** RT-PCR assays conducted on a stored saliva sample achieved similar performance to those on NP swabs and this may provide a very effective tool for population screening and diagnosis. Collection of saliva in a stabilizing solution makes the test more convenient and widely available; furthermore, the denaturing properties of the solution reduce the infective risks belonging to sample manipulation.

## INTRODUCTION

The Coronavirus Disease 19 (COVID-19) pandemic driven by the novel Severe Acute Respiratory Syndrome-CoronaVirus-2 (SARS-CoV-2) is an aggressive and potentially deadly disease firstly appeared in Wuhan, China, in 2019 and then rapidly spread worldwide (1, 2).

SARS-CoV-2 is transmitted by exposure to infectious respiratory fluids (2–6). The early detection of infected subjects plays a crucial role in their isolation to stop the spread of the infection (3, 6). Adequate sample collection and transportation are important elements for the laboratory diagnosis of SARS-CoV-2 (7). A specimen that is not collected properly may be an explanation for false negative results, as in the case of nasopharyngeal swab sampling (5, 8).

The reverse transcription polymerase chain reaction (RT-PCR) in naso-pharyngeal (NP) swabs samples is the current gold standard in detection of SARS-CoV-2 infection (5, 9, 10).

Specimen collection currently required specialized staff, exposes personal to a high risk of infection and causes discomfort to patients (11, 12). Furthermore, there is a need for a simpler and less invasive methods to make it a mass test (13–15, 15–17). An additional limiting factor, is the inadequate sampling that may result in both lower specimen quantity and in lower test sensitivity (5, 18).

To overcome these limitation, other types of biological matrices are currently under investigation (7, 18). Among these, an easily and simple-to-use alternative specimen for the detection of SARS-CoV-2 in the diagnosis of COVID-19 are saliva samples (5, 15, 19). Saliva represents a wide and homogenous resource for genomic information, useful for studying the disease status (19–22).

COVID-19 diagnostic on saliva samples overcomes the need for a trained technician and also reduces the potential risk of infection (13, 20, 21, 23). Benefits of the saliva testing include the non-invasiveness, the practical storage and the easier sample collection in particular fragile patient populations (e.g., paediatric, non-collaborative and elderly patients) (22–24). Overall, saliva testing can reduce infection spread, costs and can provide an easier access for patients (6, 25).

Considering all benefits, researchers and clinicians concur that the diagnosis and prevention of COVID-19 disease using human saliva is to be better explored (5). The non-invasive nature and ease of collection have made saliva the fluid of choice not only for diagnostic but also for more important health surveillance purposes (13, 20, 26,). Saliva is suggested to cause spread via droplet infection, appearing to be more transmissible (17, 17, 27). For example, novel investigations on saliva samples were recently proposed to understand SARS-CoV-2 variants spread (19, 22, 24, 28).

Evidences on the use of saliva, for the detection of SARS-CoV-2, are exponentially increasing, representing saliva a very attractive, readily available and repeatable biological matrix (20, 28). RT-PCR assays using saliva swab have been approved in the United States and in Japan (7, 11). In the US, the FDA has already approved saliva-based collection methods, especially in emergencies (4).

Several papers have been published on the possible use of saliva as an alternative diagnostic tool to identify SARS-COV-2 infection (19, 24, 28). The comparison of saliva and NP molecular tests has shown discrepancies between studies (15, 18, 29). In some studies, nasopharyngeal swab samples showed a greater sensitivity, others found contrary results, while in some cases saliva has proved equally effective as a diagnostic specimen (16, 18, 26, 29, 30).

Prior works have taken into question the need of stabilization buffers for collecting saliva samples, considering that the enzymatic composition could represent a negative factor for nucleic acid stabilization and protease inhibition (31). Saliva is stickiness and difficult to stabilize due to its muco-proteins and enzymatic composition (31, 32). In particular, its endogenous nucleases primarily contribute to the RNA degradation and lead to varying degrees of interference during the analytical steps, such as nucleic acid extraction and amplification by PCR (20, 29). Furthermore, saliva viscosity may reduce sample’s recovery affecting downstream applications (20, 31). Saliva collection techniques and timing, conditioning and delays in processing raw saliva samples represent the major causes for failure to apply saliva as a validated diagnostic source (20, 31, 33, 34).

Moreover, despite the hot topic, no systematic studies are available yet, in particular concerning saliva collection and analysis validation based on both patients and asymptomatic or healthy groups.

Aim of this study was to analyse the feasibility of saliva as an alternative diagnostic specimen for detecting SARS-CoV-2 infection both in patients with confirmed diagnosis of COVID-19 and in healthy people who had to undergo screening tests. The objectives were: i. to establish and validate a standardized and reproducible saliva collection method, using a saline-based solution, in order to produce high-quality RNA to improve the performance of downstream molecular investigations; ii. to compare results of the RT PCR-based molecular test on saliva samples versus NP swabs, in terms of concordance, clinical sensitivity and specificity; iii. to investigate potential association between test results and COVID-19.

## MATERIALS AND METHODS

### Study Design and Participants

In this observational study were enrolled a total of 1161 participants (age ≥18), between October 2020 and May 2021. Among these, 156 were patients with confirmed diagnosis of COVID-19 (cases), admitted to the Infectious Disease Clinic of Udine, and 1005 were asymptomatic people who had to undergo screening tests (controls). Both groups were tested for the detection of SARS-CoV-2 with NP swab and in saliva specimens. Aim of the study was to compare results of the RT PCR-based molecular test in detecting SARS-CoV-2 RNA on saliva samples and on the recommended NP swab, which is considered the gold standard. For hospitalized patients, we also looked for any correlation between the positivity of the two tests (in terms of Cycle threshold) and clinical severity, defined as a composite outcome of admission to Intensive Care Unit (ICU) or death.

All samples collected were anonymized using an alpha-numeric identification code, and the study was approved by the Local Ethics Committee.

### Naso-pharyngeal and saliva sample collection

NP specimens were collected by mid-turbinate swabbing of both nares and the posterior pharynx, avoiding the tongue. A flocked swab (*ESwab Copan*) was used for the collection of all NP clinical samples and handled as recommended in international guidelines (1, 4, 7). For saliva collection a non-invasive method was executed by making self-collected specimen in a tube container. The conserving solution contained in the saliva collection tubes was provided by Biofarma Srl and it is protected by intellectual property. The specific saline composition and the pH value made this solution optimal for the stabilization and conservation of viral RNA in saliva. Furthermore, the high saline concentration takes a denaturing effect on protein and as consequence on cells structure and proliferation. For these reasons, when saliva comes into contact with the solution, proteins and cells are disrupted hindering the viral activity. Hence, saliva samples can be considered safer for the healthcare personnel.

The subjects enrolled simply spat into the container with the conserving solution and were not asked to perform oral hygiene, to collect the sample at particular times of the day or between meals.

### Viral RNA extraction

Viral RNA was extracted and purified starting from NP swabs and saliva samples using the Virus/Pathogen Kit (*Qiagen*), on an automated Qiasymphony extractor, according to the manufacturer’s instruction. The purification procedure is designed to ensure safe, reproducible handling of potentially infectious samples and comprises three steps: lyse, bind, wash, taking the advantage of a magnetic beads principle. RNA extracted was finally eluted into a multiwall plate in a volume of 60uL of Buffer AE (*Qiagen*).

### One step Real time PCR assay

A reverse transcriptase, one-step real time PCR was employed for the detection of SARS-CoV-2 Envelope viral protein (coded by the *E-gene*). Primer sequences were selected according CDC’s guidelines and purchased from Roche (*Roche*). The amplification was performed using LightCycler® Multiplex RNA Virus Master *(Roche*), following the company’s instructions. For each reaction 5uL of template was loaded. For each reaction, a synthetic RNA positive control and a no template control were used. RT-PCR and analyses were performed employing the LightCycler 480 *(Roche*) instrument. For each amplification reaction Cp values were computed. We considered test results as positive when Cycle threshold (Ct) values were <36; according to the assay Limit of Detection (LoD) defined.

### Assay validation

The evaluation of assay accuracy was performed by testing 7 different dilutions of a patient positive sample (Cp=29), with 10 replicates each. Serial dilutions (from 1,00+E^6^ to 2 copies) of a synthetic RNA control were also computed in order to defined the expected concentration values. The precision of our analytical procedure was established as repeatability and intermediate precision and examined using the one-way ANOVA, according to UNI ISO 5725-2:2020 guidelines.

The linear range of RT-PCR was established with a series of 7-step dilutions, tested in 10 replicates each. Assay LoD was determined by utilizing different dilution of samples with known Cp values. The performance of saline collecting solution was assessed on 25 samples of saliva, matched with the fresh saliva samples collected from the same patients. The stability of saliva samples collected in the saline solution was tested at 24 and 48 hours, at two different storage temperature (room temperature RT, 18-25°C and 4°C).

### Statistics

Sensitivity, specificity, positive and negative predictive values and their 95 % confidence intervals (CI) were calculated to assess diagnostic performance. Kappa coefficient was presented to estimate agreement between swab and saliva RT-PCR test results, with its 95 % CI.

Bland-Altman analysis was used to compare the Ct values between swab and saliva results.

Descriptive statistics for categorical variables are presented as number (percent) and for continuous variables as mean ±standard deviation (SD) or median (interquartile range; IQR). Normality was assessed using Shapiro-Wilk test. Comparisons between categorical variables were performed using the Chi-square or Exact Fisher test, as appropriate. For continuous variables comparisons between two groups were done using the t-test or Mann-Whitney U test, comparisons among groups were done using ANOVA or Kruskal-Wallis test, as appropriate. For multiple comparisons, Bonferroni’s correction was applied.

For hospitalized patients, Cox regression was used to estimate association between death or admission to ICU and swab Ct, after the assumptions were verified; the same analysis was performed for saliva Ct. Performance of the models was evaluated by Harrell’s concordance index (C-index). All analyses were performed by STATA 16 statistical software and statistical significance was set at *p < 0*.*05*.

## RESULTS

### Population and diagnostic performances of the tests

In this study were enrolled a total of 1161 participants who provided both NP swab and saliva specimens for the molecular detection of SARS-CoV-2.

Among these, 156 (13.4%) were patients admitted to the Infectious Disease Clinic with confirmed diagnosis of COVID-19 (cases), while 1005 (86,6%) were asymptomatic people who had to undergo screening tests (controls).

The overall prevalence for SARS-CoV-2 molecular detection determined on NP swab was 8.9% (103/1161, 95% IC 7.3-10.7) while on saliva specimen was 10.4% (121/1161, 95% IC 8.7-12.3).

Among the 156 patients certainly affected by COVID-19 (cases) an average of 12 days (IQR 9-16 days) has passed between the onset of symptoms and the test performed for the purpose of comparison saliva to NP swab. 62.2% of cases (97/156) showed a SARS-CoV-2 positive NP swab while 71.8% (112/156) resulted positive if tested on saliva specimen. In the group of people who had to undergo screening tests (controls) the positivity with the classical swabs and with saliva specimen was reached in 0.6% (6/1005) and in 0.9% (9/1005), respectively. All these data are summarized in **Table 1**.

**Table 1:** Results of the RT PCR-based molecular test in detecting presence of SARS-CoV-2 RNA on simultaneous collection NP swab and saliva samples.

|  | NP swabs negative<br>(n = 1058) | NP swabs positive<br>(n =103) | Saliva samples<br>negative<br>(n= 1040) | Saliva samples<br>positive<br>(n=121) |
| --- | --- | --- | --- | --- |
| **Cases\***<br>(n=156) | 59/156 (37.2%) | 97/156 (62.2%) | 44/156 (28.2%) | 112/156 (71.8%) |
| **Controls\*\***<br>(n=1005) | 999/1005 (99.4%) | 6/1005 (0.6%) | 996/1005 (99.1%) | 9/1005 (0.9%) |

\*Cases = patients admitted to the Infectious Disease Clinic with confirmed diagnosis of COVID-19 (positive NP swab with consistent clinical and radiological findings). \*\*Controls = asymptomatic people who had to undergo screening tests.

Using the NP swab as the reference gold standard, the clinical sensitivity and specificity reached by saliva was 95.1% (95% CI 89–98.4%) and 97.8% (95% CI 96.8–98.6%), respectively. The Positive Predictive Value (PPV) was 81% (95% CI 72.9–87.6%) while the Negative Predictive Value (NPV) was 99.5% (95% CI 98.9–99.8%). Analyses of concordance was conducted comparing results obtained from saliva samples and the NP gold-standard swab. The agreement between the two tests was 97.6% with a Cohen’s Kappa coefficient of 0.862 (95% CI 0.812–0.912).

Concordance between tests was observed to be associated with subject’s age: in the concordant group the medium age was 50 years (IQR 30-59), while in the discordant was 70 years (IQR 61-76) with a significant difference between the two groups (*p<0*.*001*). **Table 2** shows demographic and clinical characteristics of patients with confirmed diagnosis of COVID-19 (cases).

**Table 2:** Demographic and clinical features of the case group.

|  | **Patients**<br>(n=156) |
| --- | --- |
| **Age**, median (IQR) | 72 (62-77) |
| **Days of symptoms before test**, median (IQR) | 12 (9-16) |
| **Admission to ICU**, n (%) | 34 (21.8) |
| **Death**, n (%) | 29 (18.6) |
| **Admission to ICU or death**, n (%) | 47 (30.1) |

In particular, the comparison between group with both saliva and swab concordant test (both positive or both negative) result to be significant (*p=0*.*006*; Bonferroni’s correction was applied).

Concordance of RT-PCR results was also associate with onset of symptoms before test (days) (*p=0*.*0036*), as shown in **Table 3**.

**Table 3.**
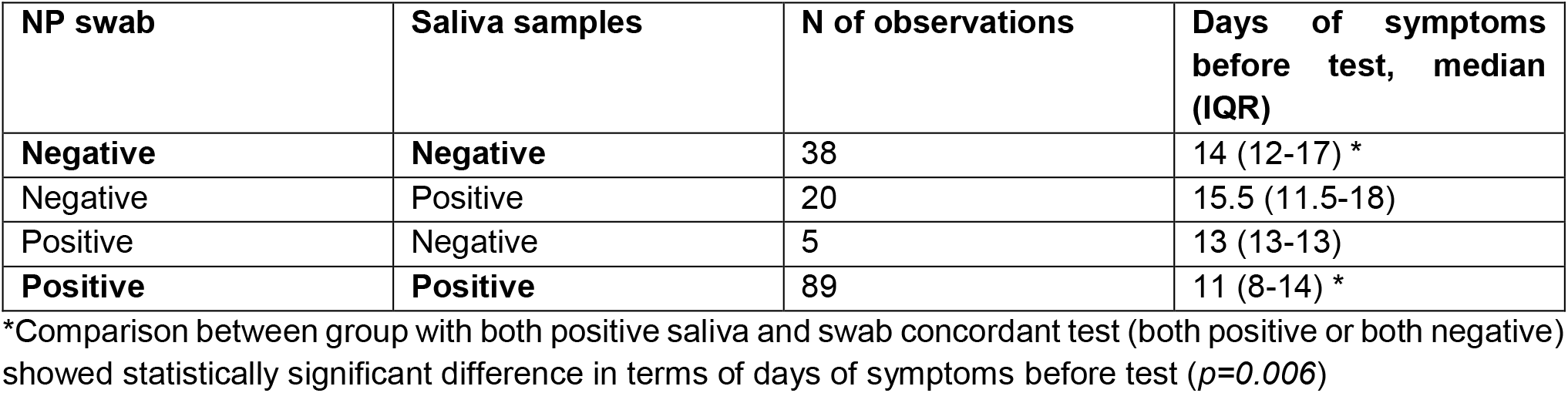
Relationship between test agreement and onset of symptoms before test (days)

| NP swab | Saliva samples | N of observations | Days of symptoms<br>before test, median<br>(IQR) |
| --- | --- | --- | --- |
| **Negative** | **Negative** | 38 | 14 (12-17) * |
| Negative | Positive | 20 | 15.5 (11.5-18) |
| Positive | Negative | 5 | 13 (13-13) |
| **Positive** | **Positive** | 89 | 11 (8-14) * |

\*Comparison between group with both positive saliva and swab concordant test (both positive or both negative) showed statistically significant difference in terms of days of symptoms before test ( $p=0.006$ )

Data regarding Ct values were available for 112/156 molecular tests based on saliva and for 71/156 NP swabs. Comparison of Ct values between saliva and swab in this group of patients showed no significant differences. The matching among positive samples is shown in **Figure 1**, where Ct values have been paired by patients.

**Figure 1:**
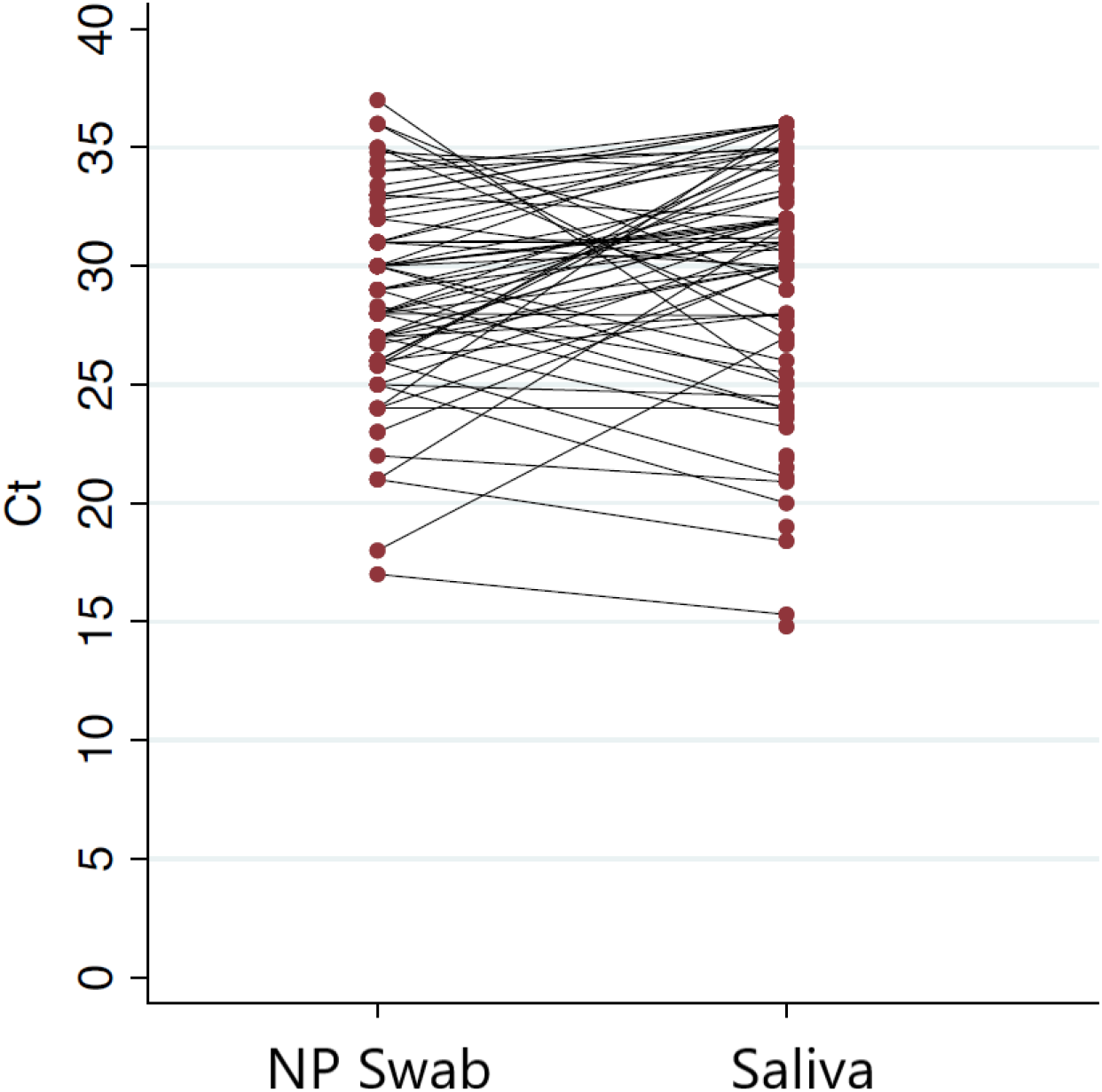
Ct values for paired samples in the 156 patients tested for SARS-CoV-2 infection.

Taking the advantage of the Bland-Altman (B&A) plot, the relationship between the Ct average value reached by the 2 tests, (x axis) and their difference (y axis) was explored, in order to assess methods’ concordance. Data presented were randomly distributed in the plot between the upper and lower limit of agreement (**Figure 2**).

**Figure 2:**
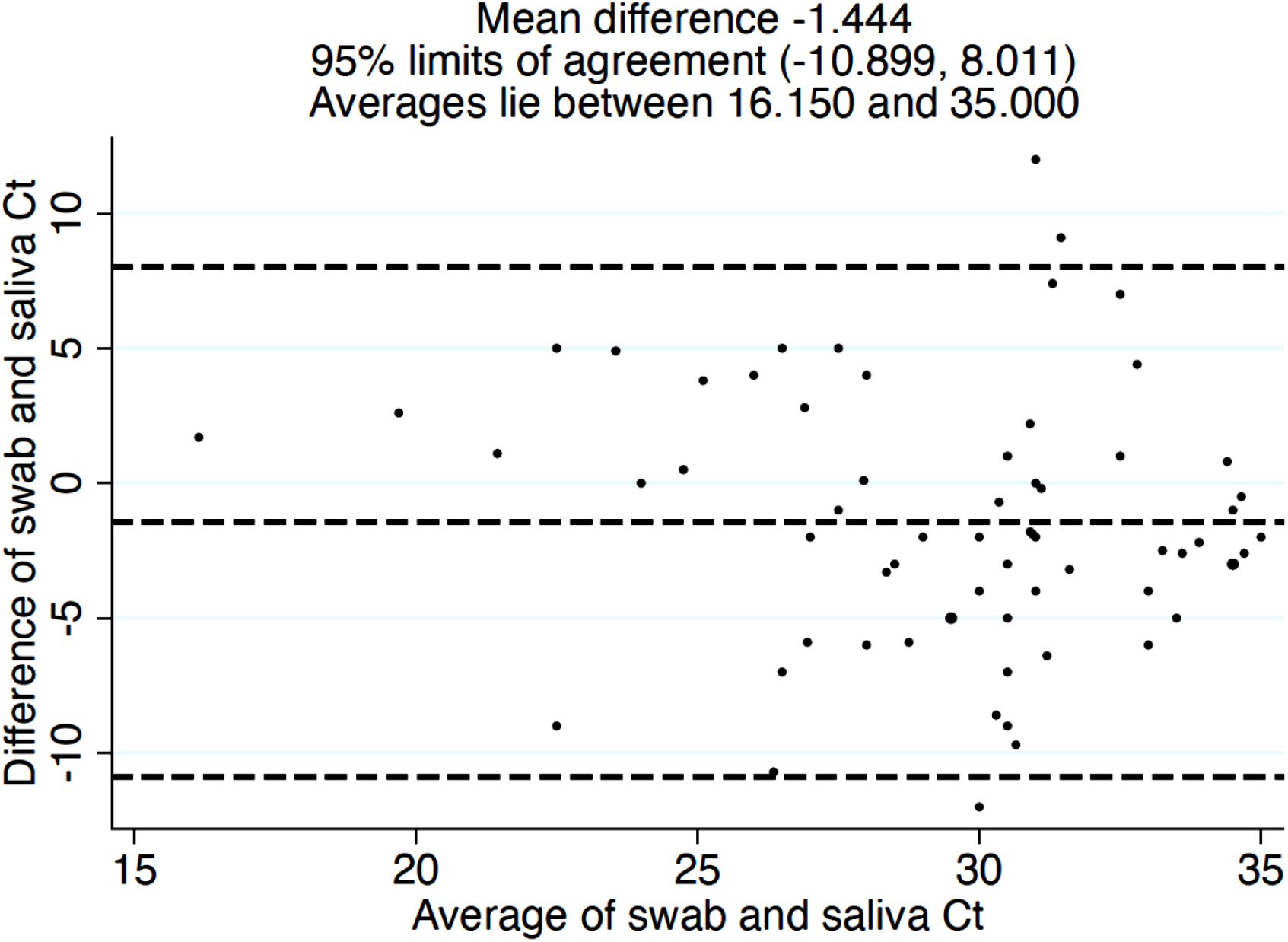
Bland-Altman (B&A) plot showing concordance between Ct values available for the 71 paired tests.

**Figure 3:**
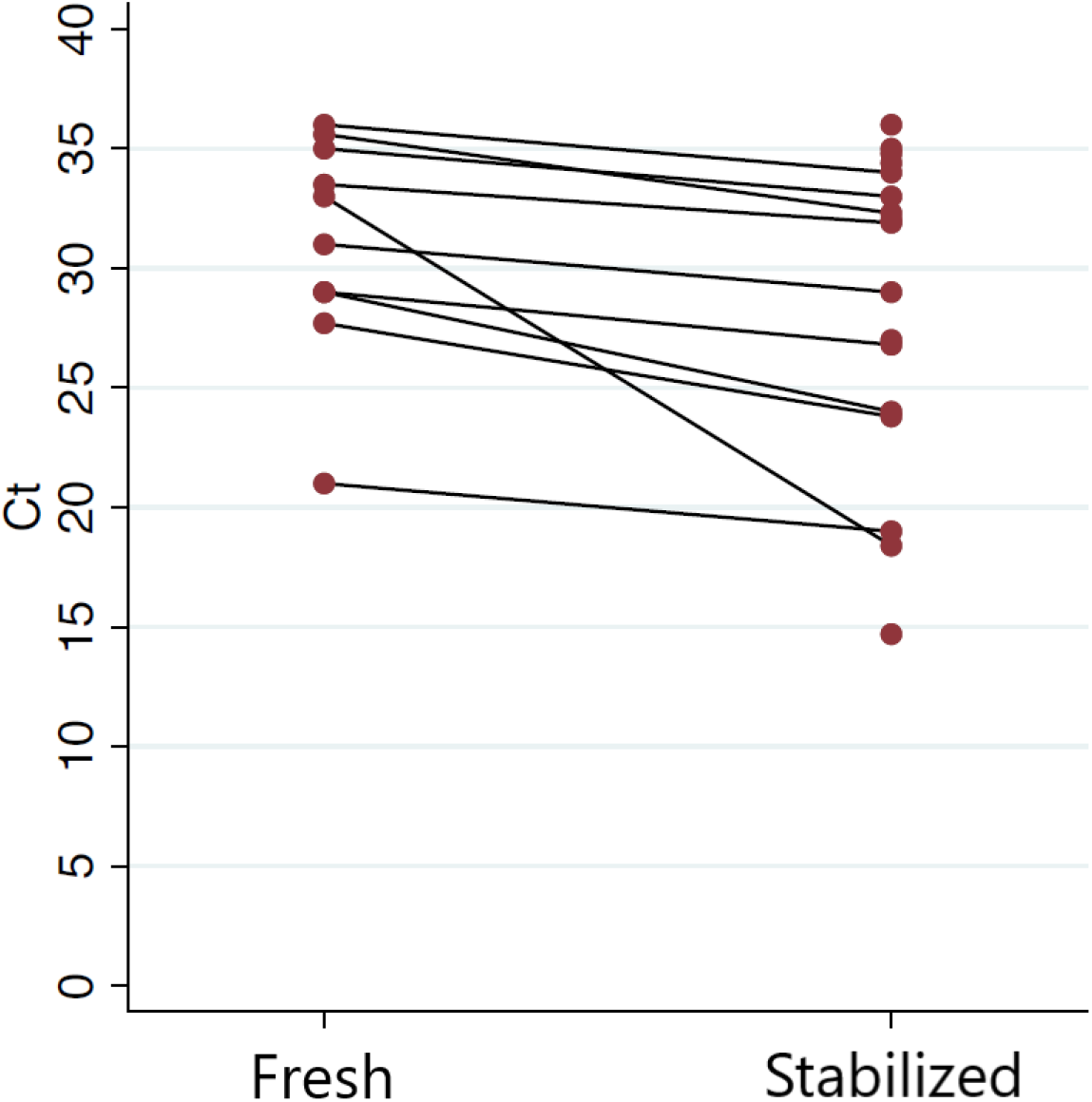
Comparison between Ct values reached from fresh and stabilized saliva samples collected from the same patients.

As regard data on Ct values, where available in both salivary and NP swabs test (n = 71 cases), we evaluated if the values found in the two tests were associated with clinical severity (composite outcome considered as orotracheal intubation or death).

The RT-PCR Ct value of both salivary and NP swab tests results to be significantly associated with COVID-19 severity in both tests as shown in **Table 4. I**n particular, we found that higher Ct values, meaning very low levels of viral RNA load found, were a protective factor.

**Table 4.** Relationship using Cox regression between Ct values where available in both saliva and NP swabs molecular test (n = 71 cases) and COVID-19 severity (composite outcome considered as orotracheal intubation or death)

|  | HR | 95% IC | p-value | Harrell's C | 95% IC |
| --- | --- | --- | --- | --- | --- |
| Ct values on NP swabs tests | 0.81 | 0.73-0.89 | **<0.001** | 0.738 | 0.628-0.848 |
| Ct values on saliva samples | 0.89 | 0.83-0.96 | **0.004** | 0.659 | 0.544-0.774 |

Furthermore, the Harrell’s C showed a better performance of the model in the case of the NP swab test (0.738 vs 0.659).

### Pre-analytical considerations: stabilizing properties of the saline solution

Saliva collected in the saline solution from 25 patients was used to preliminary evaluate its conserving properties. Comparing RT-PCR results obtained from both fresh and preserved saliva samples of a same patient, a gain of 28% positive tests (N=7/25) was observed. Considering all paired results, a significant difference between Ct values emerged depending on the collection method. In particular, stabilized saliva samples showed significantly lower Ct *(p=0*.*006)*, with a median value of 27.9 (IQR 23.8-32.3) compared with results computed from fresh samples (median Ct 32, IQR 29-35).

Furthermore, the temporal stability of SARS-CoV-2 RNA collected in the preserving saline solution, measured at different holding temperatures (20-25°C and 4°C), was about 48 hours. We found that SARS-CoV-2 viral load was reliably detected at similar levels regardless of the holding time and temperatures tested. We observed a 100% of sensitivity both at 24 and 48 hours, independently from storage temperature as confirmation that SARS-CoV-2 RNA was relatively stable. No positive sample lost were noticed. Our data indicates that viral RNA remains protected even at room temperature and at 4°C up to 48h if collected in saline medium.

### Validation of the analytical procedure

The accuracy of the RT-PCR method was measured by the coefficient of variation (CV) between measurements obtained from 7 different dilutions of a positive sample (Ct=29), with 10 replicates each. CV values reached were always less than the 20% for all the dilutions tested, which is considered a high degree of accuracy (ISO 3534-1, ISO 5725-1).

The method’s precision was evaluated based on intra-assay variation and intermediate precision (inter-assay variation), expressed as standard deviation (σ) or Relative Variation Coefficient (RDS) and derived from differences between measurements (Ct) means. Furthermore, RT-PCR analysis was repeated on 3 different days to determine the inter-day precision. Results obtained always revealed RDS values below the 5%, demonstrating a high level of precision (UNI ISO 5725-2:2020; 3534-1:2020).

Linearity was determined by serial dilutions of standard samples with known viral copies (synthetic viral sequence) in order to define reportable range. The curve plot of 8 serial dilutions (from 1,00+E^6^ to 2 copies) displayed a good linear range estimated as a goodness-of-fit (R^2^) equal to 0.996, indicating a good linear relationship between expected and observed results.

The LoD of the assay for the E-gene at 95% confidence was 5 copies per reaction, which corresponded to a Ct value of 36.15±0.51.

## DISCUSSION AND CONCLUSIONS

RT-PCR performed on NP swabs is still considered the gold standard for the diagnosis of SARS-CoV-2 infection (1, 2, 8, 11). However, the collection of NP swabs is invasive, ideally requires experience and clear instructions and has a risk of viral transmission to the sample collector (11, 14, 23). There is a need to prioritize the test for people with symptoms of COVID-19 and contacts of those with confirmed SARS-CoV-2 infection, but also to people at increased risk for exposure (for example health care workers) (2, 6); furthermore there are economic needs as well as the reopening of schools maybe they will put a strain on testing systems (6). The presence of infectious SARS-CoV-2 virus in saliva in asymptomatic or pre-symptomatic individuals has been demonstrated (9, 24, 34, 35), and this support the applicability of saliva as sample material for COVID-19 testing (15, 22). SARS-CoV-2 can be detected in the saliva of COVID-19 patients (17, 24, 28), studies confirm that saliva is effective for the identification of the SARS-CoV-2 and shows higher concentration of RNA viral copies than nasopharyngeal swabs in the same individuals (17, 22, 26, 28).

Saliva samples can sometimes be difficult to handle with existing RNA extraction methods and equipment and the heterogeneity of saliva specimen can represent limitations, but there are several advantages (14, 31, 32, 36). Actually, saliva sample collection is non-invasive and can be easily performed by the individual themselves and this could reduce the risk of transmission to the sample takers (13, 14, 17). In our study, a high-concentrate saline solution was employed to collect saliva samples. The ionic strength of the solution acts as denaturing agent and destroys all protein and muco-protein’s structure, interfering on the interaction between the virus and the host cell. This observation represents an additional key aspect in mitigate the risk of infection during sample collection and allows to bypass the absence of dedicate spaces for the safe conduction of laboratory activity. In this work, the combination of the saliva self-collection procedure with the inhibiting property of Bsawb solution, could be considered an advancement to facilitate COVID-19 diagnostics.

Furthermore, saliva collection facilitates sampling of children or disabled and this can increase the acceptance to routine testing practices performed at repeated intervals among high-risk populations. This modality of sample collection is easy and non-invasive, so it may be performed by non-healthcare professionals or individuals themselves who are properly instructed (13, 14, 16, 35).

Overall, recent studies suggest that the diagnostic sensitivity of RT-PCR on saliva samples is variable (35), often lower but sometimes higher than that of NP swabs, and sensitivity varies when considering different saliva collection technique (29, 37). In fact, this variability is often related to an inappropriate saliva sampling (13, 14, 36, 38). In our study the collection of saliva in a saline solution able to inactivate proteases allowed to recover false negative cases derived from an incorrect pre-analytical sampling by NP swab. Saliva degrades very quickly and this could result in an higher quantity of negative tests, affecting test sensitivity (31, 38, 39). The constituents of saliva can significantly affect the quality of viral RNA indeed. RNA is very labile and sensitive to the degradation caused by RNAse and endonuclease activity as well as temperature, which can reduce RNA concentration available for the molecular downstream applications (12, 31). Improving the performance of the current saliva collection procedures had the advantage to obtain high-quality RNA for downstream RT-PCR test (14, 36, 36). Furthermore, the stability of nucleic acids in the saline solution guarantees the quality of salivary RNA also as stored at room temperature for 48 hours.

All these advantages, allowed to validate the SARS-CoV-2 molecular test on saliva samples in a cohort of 1161 subjects, composed by both patients and controls.

Our experience demonstrated that the clinical sensitivity and specificity reached by test on saliva was 95.1% (95% CI 89–98.4%) and 97.8% (95% CI 96.8–98.6%), respectively, referring to NP swab as gold standard and that agreement between the two tests was very high (97.6%, Cohen’s Kappa coefficient of 0.862, 95% CI 0.812–0.912).

Our data shown also that concordance between tests was observed to be associated with subject’s age and that in the discordant was 70 years (IQR 61-76) with a significant difference (*p<0*.*001*), but this is because the control group was made up of younger and healthier people individuals who underwent screening tests, while the cases were patients hospitalized due to COVID-19.

Taking into account all data, with this study we demonstrated that saliva could be interchangeably and equally applicable for the identification of SARS-CoV-2 infection, for both diagnostic and screening intents.

In the study by Silva et al., not yet peer-reviewed and published as a preprint, saliva viral load seems to correlate with a spectrum of disease severity throughout the course of illness and it appears as a predictor of mortality(8)

In our work, we found that COVID-19 severity (composite outcome considered as admission to ICU or death) was correlated with the Ct values found in both saliva and NP swabs: higher Ct values were considered a protective, meaning that very low levels of viral RNA were found. This analysis showed slightly better performance for the NP swabs and this could be attributable to a faster viral load clearance in NP swabs than in saliva specimen, which with the high sensitivity demonstrated in this work, it can show a prolonged positivity for viral RNA.

This study, however, was conducted in a period in which the epidemic in our region was sustained by the wild-type strain: it is possible that with the arrival of variants that compartmentalize more in saliva, the results may differ from those found and show better performance for saliva in predicting clinical outcome (9,40,41)

In conclusion, we demonstrated the diagnostic value of saliva as an alternative matrix for SARS-CoV-2 molecular detection. RT-PCR performed on saliva could be applied to the established methods and may provide an additional back-up for population screening. The collection of saliva in a conserving saline solution by patients themselves makes the test more affordable and widely available, preventing the spread of the infection. The test may also look for viral proteins to screen large numbers of asymptomatic people and in a future perspective it can be used also to characterize SAR-CoV-2 genetic variants, which are considered more transmissible for their compartmentalization in saliva, their spread and high replication rate.

## Data Availability

The authors confirm that all relevant data are included in the article and materials are available on request.

## ABBREVIATIONS

COVID-19: Coronavirus Disease 19
SARS-CoV-2: Severe Acute Respiratory Syndrome-CoronaVirus-2
NP: Nasopharyngeal Swab
RT-PCR: Reverse transcription polymerase chain reaction
PPV: Positive Predicted Value
NPV: Negative Predicted Value
Ct: Cycle threshold
ICU: Intensive Care Unit
LoD: Limit of Detection
RT: Room Temperature
CI: Confidence Intervals
SD: Standard Deviation
IQR: Interquartile Range
HR: Hazard Ratio

## ACKNOWLEDGMENTS

The authors are thankful to Biofarma Srl to provide the collection devices employed in the study.

## AUTHORS’ CONTRIBUTIONS

MB optimized and performed the experimental analysis and drafted the manuscript; ES coordinated the patient recruitment, collected clinical samples, created and updated the clinical database; helped to draft the manuscript; BM participated to the experiment design and helped in method’s validation; MDM performed the statistical analysis; DC participated in the analysis and in data interpretation; CDC, EV, AZF and JDP collected clinical samples and updated the clinical database; RM and AM helped in clinical specimen processing; EN helped in the experimental set up, reviewed the manuscript; CT coordinated the patient recruitment and supervised the study; draft the manuscript; MI designed the study and performed the statistical analysis; FC conceived, supervised and coordinated the study; draft the manuscript.

All authors reviewed and approved the final manuscript.

## COMPETING INTERESTS

the authors declare that they have no competing interests.

## FUNDING

This research was funded by PRIN 2017 n.20178S4EK9 ‒ “Innovative statistical methods in biomedical research on biomarkers: from their identification to their use in clinical practice”.

## Notes

### Competing Interest Statement

The authors have declared no competing interest.

### Author Declarations

Ethical approval was obtained from the Medical Research Ethics Committee of the Region Friuli Venezia Giulia, Italy. Consent CEUR-2021-OS-14

